# Remdesivir in treatment of COVID-19: A systematic benefit-risk assessment

**DOI:** 10.1101/2020.05.07.20093898

**Authors:** Miranda Davies, Vicki Osborne, Samantha Lane, Debabrata Roy, Sandeep Dhanda, Alison Evans, Saad Shakir

**Affiliations:** Drug Safety Research Unit, Southampton, UK; School of Pharmacy and Biomedical Sciences, University of Portsmouth, Portsmouth, UK

## Abstract

**Background:** There is a need to identify effective, safe treatments for COVID-19 (coronavirus disease) rapidly, given the current, ongoing pandemic. A systematic benefit-risk assessment was designed and conducted to strengthen the ongoing understanding of the benefit-risk balance for remdesivir in COVID-19 treatment by using a structured method which uses all available data.

**Methods:** The Benefit-Risk Action Team (BRAT) framework was used to assess the overall benefit-risk of the use of remdesivir as a treatment for COVID-19 compared to standard of care, placebo or other treatments. We searched PubMed,Google Scholar and government agency websites to identify literature reporting clinical outcomes in patients taking remdesivir for COVID-19. A value tree was constructed and key benefits and risks were ranked by two clinicians in order of considered importance.

**Results:** Several key benefits and risks for use of remdesivir in COVID-19 compared to placebo have been identified. In one trial, the benefit of time to clinical improvement was not statistically significant (21 vs 23 days, HR=1.23, 95% CI: 0.87, 1.75), although the study was underpowered. In another trial, a shorter time to recovery in patients treated with remdesivir was observed (11 vs 15 days), with non-significant reduced mortality risk (8% vs 12%). Risk data were only available from one trial. This trial reported fewer serious adverse events in patients taking remdesivir (18%) comparted to the placebo group (26%), however more patients in the remdesivir group discontinued treatment as a result of an adverse event compared to those patients receiving placebo (12% vs 5%).

**Conclusions:** Preliminary clinical trial results suggest a favourable benefit-risk profile for remdesivir compared to placebo, however there is limited safety data available at the current time. The current framework summarises the key anticipated benefits and risks for which further data are needed. Ongoing clinical trial data can be incorporated into the framework when available to provide an updated benefit-risk assessment.

## 1 Introduction

Coronaviruses are a cause of respiratory tract infections in humans [1]. A novel coronavirus emerged in Wuhan, China in December 2019 [2], subsequently called Severe Acute Respiratory Syndrome coronavirus 2 (SARS-CoV-2) [3]. Coronavirus disease (COVID-19) is caused by SARS-CoV-2 [3] and since March 2020 this outbreak has been declared a pandemic by the World Health Organisation (WHO) [4]. Fever, cough and shortness of breath are the main reported symptoms of COVID-19 [5] but it also has a concerning case mortality rate among certain populations, such as older adults and those with underlying health conditions.

Currently, there is a need to identify effective, safe treatments for COVID-19 as quickly as possible, due to the ongoing pandemic. One such proposed treatment is remdesivir, which is an investigational antiviral medicine with proven activity against RNA viruses [6, 7]. Remdesivir was originally developed for treatment of Ebola Virus Disease and results from the phase 3 randomised clinical trial for this indication have been published [8]. It has been shown to have broad spectrum in-vitro activity against several coronaviruses, including SARS-CoV-2 [6, 9, 10] in-vivo activity against MERS-CoV in animal models [11, 12], and has been made available for patients with COVID-19 through compassionate use programmes [13, 14]. Several clinical trials are ongoing to examine the effectiveness and safety of remdesivir in COVID-19 in humans [15-18]. Most recently, remdesivir was granted an Emergency Use Authorisation in COVID-19 by the US Food and Drug Administration. Whilst there is considerable interest in the use of Remdesivir for COVID-19, there has not been a systematic benefit-risk assessment on the use of remdesivir for COVID-19 treatment using a structured descriptive framework.

The ongoing monitoring of the benefit-risk balance for remdesivir in COVID-19 treatment is strengthened by use of a systematic assessment. The Benefit-Risk Action Team (BRAT) framework allows identification of the key benefits and risks of a product in a defined disease context within a structured descriptive framework; further quantitative assessments can then be applied and conducted according to the availability of relevant data at that time [19]. The BRAT framework was also specifically designed to assist communication with regulatory authorities [20]. The decision-making process is transparent due to the framework design, while any assumptions can be explored further by sensitivity analysis through a quantitative component [21].

This benefit-risk assessment has been conducted based on publicly available publications to date (data-lock April 30th 2020). It is however acknowledged that there is extremely limited data available at this timepoint. The benefit-risk assessment has been designed to be implemented regardless of the quantity of data available, but the intention is that the framework will subsequently be readily available to repeat the assessment as further data arise, e.g. results from new and ongoing clinical trials, allowing for rapid decision-making.

## 2 Objectives

To examine the benefit-risk profile of remdesivir in COVID-19 patients compared to standard of care, placebo or other treatments.

## 3 Methods

### 3.1 Benefit-Risk Framework

The BRAT framework was used to assess the overall benefit-risk of using remdesivir as a treatment for COVID-19 compared to standard of care, placebo or other treatments. BRAT uses a six step iterative process to support the decision and communication of a Benefit-Risk Assessment: define decision context, identify outcomes, identify data sources, customise framework, assess outcome importance, and display and interpret key Benefit-Risk metrics [20, 21]. We identified three settings of interest for use of COVID-19 treatments; treatment for severe disease, treatment of milder disease in the community, and prevention in health care professionals exposed to the virus. We have focused on the use of remdesivir for the treatment of severe COVID-19 disease within this benefit-risk assessment. For the purposes of this study, we considered severe COVID-19 to include any patient who was hospitalised as a result of the infection.

#### 3.1.1 Population of interest

The population of interest were patients with severe COVID-19, while the exposure of interest was remdesivir. The comparators of interest were standard of care, placebo or other treatments for COVID-19.

#### 3.1.2 Outcomes of interest

Regardless of importance, all potential benefits and risks related to remdesivir were initially identified. Key benefits and risks were then selected by clinician judgement, which were those considered to drive the benefit-risk balance of remdesivir. The key benefits and risks were used to construct a value tree, ranking benefits and risks in order of considered importance.

#### 3.1.3 Data sources and customisation of the framework

We searched PubMed, Google Scholar and government agency websites to identify suitable data for inclusion. The search strategy used was:

> remdesivir AND (coronavirus OR covid* OR "SARS-CoV-2" OR "2019-NCov")

Papers were included if they reported quantitative data on effectiveness and/or safety of remdesivir in patients with severe COVID-19. Case reports were excluded. Results were restricted to English language only (abstracts in English language were acceptable where sufficient data was provided) and peer-reviewed publications (for PubMed and Google Scholar) since 2019 to 30th April 2020. Where a control group was included, data were extracted for each benefit and risk, for both remdesivir and the comparator (standard of care, placebo or other treatments).

### 3.2 Outcome assessment

A summary benefit-risk table was created to allow visualisation of the magnitude of each benefit and risk. Risk differences and corresponding 95% confidence intervals (CI) were calculated for each outcome where both numerator (number of events) and denominator (number of patients at risk) were available for both the treatment group (remdesivir) and comparator group.

#### 3.2.1 Quantitative assessment

Due to paucity of data, a fully quantitative assessment was not undertaken. However, the risks and risk difference per 1000 patients were calculated for each benefit and risk. The outcomes identified in the value tree were ranked so that swing weighting can be applied in future assessments. The weighted net clinical benefit (wNCB) can subsequently be calculated using these weights [21-23]. We would propose using the Sutton et al method, where benefits have a positive contribution to the wNCB and risks have a negative contribution [23]; the overall wNCB would be considered positive (benefit outweighs the risk) where wNCB >0. A sensitivity analysis can also be used to examine the robustness of the assigned weights and whether significant changes would alter the benefit-risk profile [19]. Analysis using wNCB was not undertaken at this time due to the limited release of clinical trial data.

## 4 Results

Figure 1 displays the value tree of the key benefits and risks related to remdesivir treatment in COVID-19. The benefits included in the value tree include key endpoints included in clinical trial protocols for studies assessing the efficacy of remdesivir in severe COVID-19 disease. As remdesivir is currently not approved for use in any condition, it is acknowledged that its safety profile has not been completely characterised. For the purposes of identifying potentials risks associated with the use of remdesivir in COVID-19 disease, safety data has been identified from currently available sources. These include studies reporting its use in the treatment of Ebola Virus Disease [8], cases series documenting the use of remdesivir in COVID-19 disease [24], and safety data included in the study by Wang et al [25]. Both the efficacy and safety outcomes have been presented in ranked numerical order according to perceived clinical significance.

**Fig 1.**
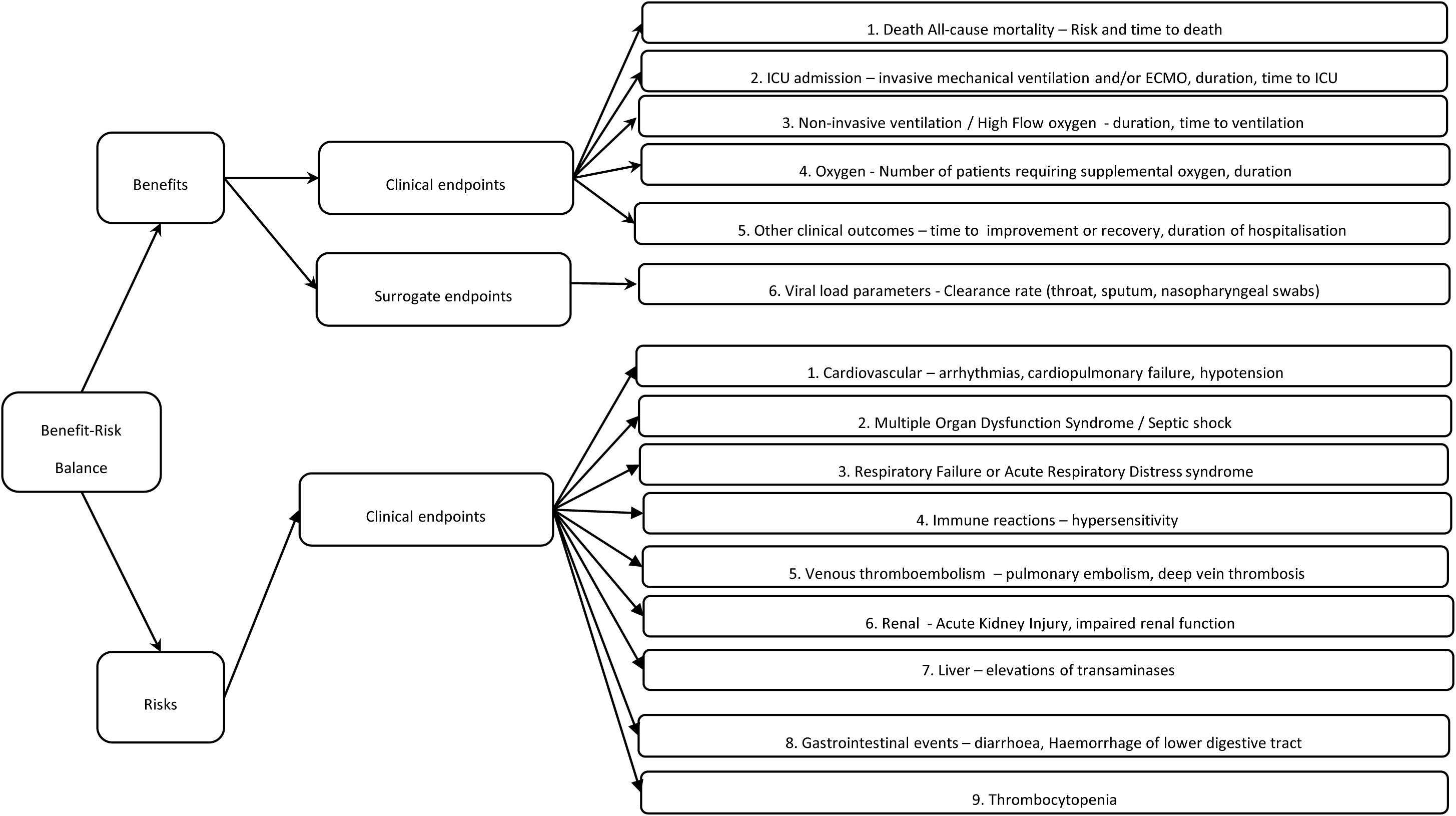
Value tree of key benefits and risks identified for remdesivir, ranked by order of clinical significance

### 4.1 Benefits

The benefits listed in the value tree represent key clinical endpoints included in clinical trial protocols. These have been ranked in order of perceived clinical importance, from the clinical endpoint of mortality risk through to the surrogate endpoint of viral clearance. Many studies have utilised ordinal scales, which include a spectrum of the clinical status of the patient. The primary endpoint used in the recently published study by Wang et al [25] was time to clinical improvement up to day 28; this was defined as the time (in days) from randomisation to the point of a decline of two levels on a six-point ordinal scale of clinical status (from 1=discharged to 6=death) or discharged alive from hospital, whichever came first. The six-point scale was as follows: death=6; hospital admission for extracorporeal membrane oxygenation or mechanical ventilation=5; hospital admission for non-invasive ventilation or high-flow oxygen therapy=4; hospital admission for oxygen therapy (but not requiring high-flow or non-invasive ventilation)=3; hospital admission but not requiring oxygen therapy=2; and discharged or having reached discharge criteria =1.

The primary endpoint in the Multicentre, Adaptive, Randomized Blinded Controlled Trial of the Safety and Efficacy of Investigational Therapeutics for the Treatment of COVID-19 in Hospitalized Adults, known as the Adaptive COVID-19 treatment trial sponsored by the National Institute of Allergy and Infectious Diseases (NIAID) [26] was time to recovery [Time Frame: Day 1 through Day 29]. Day of recovery was defined as the first day on which the subject satisfied one of the following three categories from the ordinal scale: 1) Hospitalized, not requiring supplemental oxygen - no longer requires ongoing medical care; 2) Not hospitalized, limitation on activities and/or requiring home oxygen; 3) Not hospitalized, no limitations on activities. Other key clinical outcomes included duration of ventilation, duration of oxygen support, and duration of hospital stay, in addition to time from randomization to discharge or death. Surrogate endpoints assessing viral load clearance are likely to be less robust forms of endpoint data, and accordingly have been ranked lower in terms of clinical importance.

### 4.2 Risks

As remdesivir is an unapproved medicine in most countries, its safety profile has not yet been fully characterised. Known key risks at the current time have been included in the value tree, identified from a variety of sources, including recently published clinical trial data in the context of COVID-19 [25]. These have also been ranked according to perceived seriousness.

Cardiovascular outcomes including hypotension and arrhythmias have been documented following the use of remdesivir [8, 27, 24], however the risk of cardiovascular outcomes with remdesivir remains largely unknown [28]. Hypotension was reported in one patient in the phase 3 study investigating remdesivir in the context of Ebola Virus Disease; this patient subsequently suffered a cardiac arrest, although the author states that this death could not readily be distinguishable from underlying fulminant Ebola Virus Disease itself [8]. Cardiac arrest was also reported in a further patient in the remdesivir group reported in the COVID-19 study by Wang et al [25]. Multiple organ dysfunction, septic shock, acute kidney injury and hypotension have also been reported as adverse events amongst patients provided with remdesivir either on a compassionate-use basis, or in a clinical trial [24, 25]. Respiratory failure or acute respiratory distress syndrome has been cited as an adverse event in patients taking remdesivir [25, 29], and as such has been included here, although it is acknowledged that this may be related to underlying disease (COVID-19), rather than to remdesivir. Elevations in liver transaminases and gastrointestinal events, including diarrhoea, have also been reported with the use of remdesivir [13, 8, 24, 30], in addition to reports of haemorrhage of the lower gastrointestinal (GI) tract [25, 31].

### 4.3 Quantitative data

Where available, clinical trial data relating to any efficacy or safety outcomes included in the value tree for both remdesivir and a comparator group were extracted and included in Table 1. Based on this data, summary metrics including risks per 1000 patient years, and risk differences have been presented in Table 2. We identified 68 papers through literature searching from PubMed and 384 papers from Google Scholar. We also identified results from one clinical trial on the NIAID website. After initial review, one paper and results from one clinical trial on the NIAID website were included in the final benefit-risk assessment.

**Table 1.** Data for key benefits and risks identified for remdesivir from published literature

| Outcome name | Study first author | Study primary outcome | Setting | Remdesivir risk estimate | Remdesivir number of patients | Remdesivir number of events | Comparator type | Comparator risk estimate | Comparator number of patients | Comparator number of events | RD point estimate | RD lower 95% CI | RD upper 95% CI |
| --- | --- | --- | --- | --- | --- | --- | --- | --- | --- | --- | --- | --- | --- |
| **Benefits** |  |  |  |  |  |  |  |  |  |  |  |  |  |
| Death | NIAID | Time to recovery | Hospital | 0.08 |  |  | Placebo | 0.12 |  |  | -0.04 |  |  |
| Death at day 28 | Wang et al | Time to improvement | Hospital | 0.14 | 158 | 22 | Placebo | 0.13 | 78 | 10 | 0.01 | -0.08 | 0.10 |
| ICU admission- invasive ventilation at day 28 | Wang et al | Time to improvement | Hospital | 0.01 | 150 | 2 | Placebo | 0.04 | 77 | 3 | -0.03 |  |  |
| ICU admission- duration of invasive ventilation (median days) | Wang et al | Time to improvement | Hospital | 7.0 |  |  | Placebo | 15.5 |  |  | -4.0 | -14.0 | 2.0 |
| Non-invasive ventilation at day 28 | Wang et al | Time to improvement | Hospital | 0.01 | 150 | 2 | Placebo | 0.03 | 77 | 2 | -0.02 |  |  |
| Oxygen at day 28 | Wang et al | Time to improvement | Hospital | 0.12 | 150 | 18 | Placebo | 0.17 | 77 | 13 | -0.05 |  |  |
| Duration of oxygen (median days) | Wang et al | Time to improvement | Hospital | 19.0 |  |  | Placebo | 21.0 |  |  | -2.0 | -6.0 | 1.0 |
| Time to clinical improvement (median days) | Wang et al | Time to improvement | hospital | 21.0 | 158 |  | Placebo | 23.0 | 78 |  | -2.0 |  |  |
| Improvement at day 28 | Wang et al | Time to improvement | Hospital | 0.65 | 158 | 103 | Placebo | 0.58 | 78 | 45 | 0.08 | -0.06 | 0.21 |
| Time to recovery (median days)* | NIAID | Time to recovery | Hospital | 11.0 |  |  |  | 15.0 |  |  |  |  |  |
| Viral load parameters- undetectable viral RNA at day 28 | Wang et al | Time to improvement | Hospital | 0.76 | 131 | 99 | Placebo | 0.83 | 65 | 54 | -0.08 | -0.19 | 0.04 |
| **Risks** |  |  |  |  |  |  |  |  |  |  |  |  |  |
| Cardiac arrest | Wang et al | Time to improvement | Hospital | 0.01 | 155 | 1 | Placebo | 0.00 | 78 | 0 | 0.01 |  |  |
| Cardiopulmonary failure | Wang et al | Time to improvement | Hospital | 0.05 | 155 | 8 | Placebo | 0.09 | 78 | 7 | -0.04 |  |  |
| Tachycardia | Wang et al | Time to improvement | Hospital | 0.00 | 155 | 0 | Placebo | 0.01 | 78 | 1 | -0.01 |  |  |
| Multiple organ dysfunction syndrome | Wang et al | Time to improvement | Hospital | 0.01 | 155 | 1 | Placebo | 0.03 | 78 | 2 | -0.02 |  |  |
| Septic shock | Wang et al | Time to improvement | Hospital | 0.01 | 155 | 1 | Placebo | 0.01 | 78 | 1 | 0.00 |  |  |
| Acute Respiratory Distress Syndrome | Wang et al | Time to improvement | Hospital | 0.10 | 155 | 16 | Placebo | 0.08 | 78 | 6 | 0.02 |  |  |
| Hypersensitivity | No data |  |  |  |  |  |  |  |  |  |  |  |  |
| Pulmonary Embolism | Wang et al | Time to improvement | Hospital | 0.01 | 155 | 1 | Placebo | 0.01 | 78 | 1 | -0.01 |  |  |
| Deep Vein Thrombosis | Wang et al | Time to improvement | Hospital | 0.01 | 155 | 1 | Placebo | 0.01 | 78 | 1 | -0.01 |  |  |
| Acute Kidney Injury | Wang et al | Time to improvement | Hospital | 0.01 | 155 | 1 | Placebo | 0.00 | 78 | 0 | 0.01 |  |  |
| Increased alanine aminotransferase leading to discontinuation | Wang et al | Time to improvement | Hospital | 0.01 | 155 | 2 | Placebo | 0.00 | 78 | 0 | 0.01 |  |  |
| Haemorrhage of lower digestive tract | Wang et al | Time to improvement | Hospital | 0.01 | 155 | 1 | Placebo | 0.00 | 78 | 0 | 0.01 |  |  |
| Thrombocytopenia | Wang et al | Time to improvement | Hospital | 0.01 | 155 | 1 | Placebo | 0.00 | 78 | 0 | 0.01 |  |  |
| Any AE which was reason for discontinuation | Wang et al. | Time to improvement | Hospital | 0.12 | 155 | 18 | Placebo | 0.05 | 78 | 4 | 0.07 |  |  |
| Any AE defined as serious | Wang et al. | Time to improvement | Hospital | 0.18 | 155 | 28 | Placebo | 0.26 | 78 | 20 | -0.08 |  |  |

\*=p<0.05; RD=Risk difference; CI= confidence interval; AE=Adverse Event

**Table 2.**
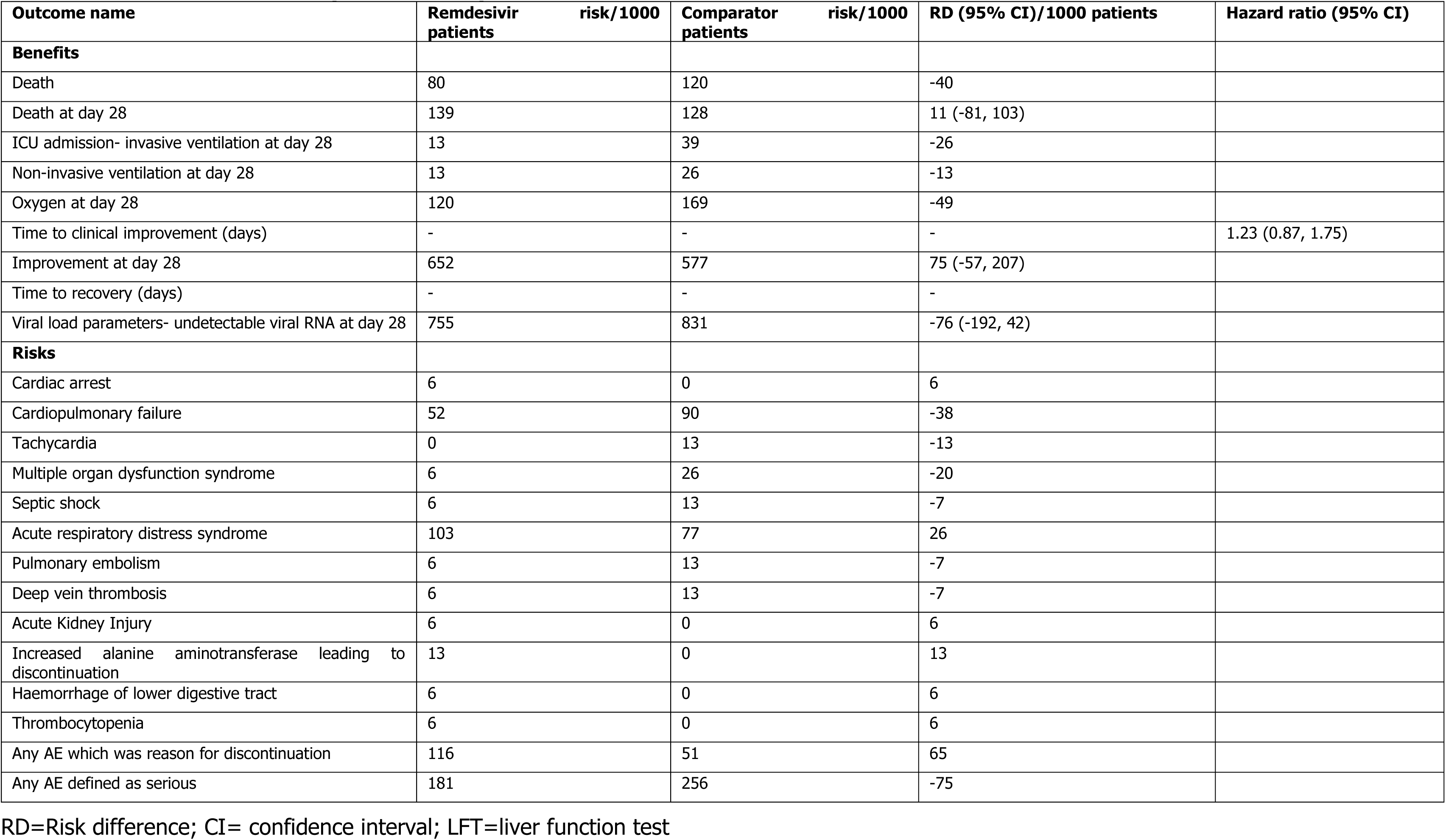
Benefit-Risk summary table for key benefits and risks identified for remdesivir

In the Wang et al trial, the benefit of time to clinical improvement was not statistically significant (21 vs 23 days, HR=1.23, 95% CI: 0.87, 1.75). In the NIAID trial, a statistically significant shorter time to recovery in patients treated with remdesivir was observed (11 vs 15 days; p<0.001), with non-significant reduced mortality risk (8% vs 12%; p=0.059). Other non-significant benefit data were identified from the Wang et al trial including invasive ventilation and oxygen use at day 28 (1% vs 4% and 12% vs 17%, respectively).

Risk data were only available from the Wang et al trial, which reported fewer serious adverse events in patients taking remdesivir (18%) comparted to the placebo group (26%), however more patients in the remdesivir group discontinued treatment as a result of an adverse event compared to those patients receiving placebo (12% vs 5%).

## 5 Discussion

This benefit-risk assessment presented the currently known key benefits and risks for the use of remdesivir for severe COVID-19 disease. The value tree provides a visual summary of these key benefits and risks, which have been ranked according to perceived clinical importance. As remdesivir is currently unlicensed in most countries, these include endpoints used in remdesivir clinical trials, and risks identified from available data sources. A literature search (including grey literature from government agency websites) identified relevant numerical data for these outcomes for both remdesivir, and comparator groups. Recently, the first results from a randomised, double-blind, placebo-controlled clinical trial studying remdesivir in the context of COVID-19 have been published [25]. The primary outcome suggested a reduction in the median time to clinical improvement, although this difference was non-significant (21 vs 23 days, HR=1.23, 95% CI: 0.87, 1.75). However, the planned sample size of 453 patients (151 on placebo and 302 on remdesivir) was not reached due to difficulties in enrolment, and therefore this study may have been underpowered to detect significant differences. Nevertheless, multiple additional endpoints were included in this study, and these data have been presented in Table 1. Whilst no statistically significant differences were observed between the remdesivir and placebo group [25], the study reported generally improved outcomes in the remdesivir group compared to placebo, including reductions in mortality, risk of invasive and non-invasive ventilation, and need for supportive oxygen at day 28.

Data has also been presented for remdesivir and the placebo group included in the Adaptive COVID-19 treatment trial [26]. As of 29^th^ April 2020, limited data has been made available on the NIH website [26] and suggested a statistically significant reduction in time to recovery from a median 15 days (placebo group) to 11 days (remdesivir group) (p<0.001). Further study results provided suggested a non-significant reduced mortality risk amongst remdesivir patients (8.0%) vs. patients given placebo (11.6%), p=0.059, which equates to 40 fewer deaths per 1000 patients treated with remdesivir.

Key risks have been identified and included based on currently available evidence, however it is acknowledged that the safety profile of remdesivir has not been fully characterized. From risk data currently available, key risks were identified and ranked according to seriousness. The cardiovascular side effects of remdesivir are largely unknown; one patient in the study by Wang et al was noted to have had a cardiac arrest. Individual cases of multiple organ dysfunction syndrome and septic shock were also reported in this study [25], whilst reports were also identified in patients receiving remdesivir on a compassion ate use basis [24]. In the latter case, reports of these events occurred in patients who were on invasive ventilation. Whilst there were multiple reports of liver enzyme abnormalities in patients who received remdesivir in the study by Wang et al [25], three patients discontinued treatment as a result of liver enzyme elevation (two reports of raised alanine aminotransferase, one report of increased total bilirubin). Reports of acute respiratory distress syndrome have also been reported as adverse events in patients taking remdesivir [25, 29]. Sixteen patients experienced this outcome during treatment in the study by Wang et al, [25], which led to discontinuation in seven patients. Finally, whilst fewer patients experienced an adverse event classified as serious in the remdesivir group (18%) compared to the placebo group (26%), a higher number of patients discontinued remdesivir as a result of an adverse event (12% vs. 5%) [25].

Currently there would appear to be favourable efficacy results from patients treated with remdesivir in the context of severe COVID-19 disease from these preliminary data available. The data in the study by Wang et al [25] did not include any statistically significant results, although it is acknowledged that the study was underpowered. Primary endpoint data available from the Adaptive COVID 19 trial [26] have also suggested a shorter time to recovery in patients treated with remdesivir, with non-significant reduced mortality risk. There is limited safety data available at the current time, although it is expected that this will increase as more clinical trial safety data becomes available. From the limited clinical trial safety data, it is unclear whether reports of serious adverse events are a function of the underlying severe COVID-19 disease, or attributable to treatment with remdesivir. It is anticipated that both the efficacy, and the safety profile will be further strengthened in the coming months with the availability of additional clinical trial data.

### 5.1 Strengths and Limitations

Sample sizes for each outcome were limited to those available in the original studies and may not have adequate power to detect differences in risk between groups, especially where the outcomes examined were not the primary outcome of interest. The benefit-risk assessment is limited by the availability of data in the published literature. However, this assessment can be subsequently updated once further data from clinical trials are available. In addition, given the public health urgency of the COVID-19 pandemic, it is important to provide a systematic assessment of the benefits and risks of remdesivir treatment with evidence available to date and establish a framework which can be used to rapidly update the assessment once further data become available.

Data quality is not reflected in this benefit-risk assessment, though all data was extracted from peer-reviewed manuscripts, with the exception of data included from the Adaptive COVID-19 treatment trial. These data were obtained from the NIAID website, and included time to recovery and mortality data. Since the full results of this clinical trial were not made available at the time of datalock (April 30^th^ 2020), a wNCB analysis was not undertaken because there was a concern that only including efficacy outcomes from the Adaptive COVID-19 treatment trial may bias the wNCB results. It is anticipated that such an analysis can be undertaken in the future when results are made available, to provide further evidence on the benefit-risk profile.

Confirmation of causality was not a requirement for inclusion of data in the BRAT assessment. Patients may have been on other concomitant medications or had other medical conditions at the time of remdesivir treatment. Finally, we considered hospitalisation of patients to reflect severe COVID-19, but we acknowledge that severity of disease may vary regardless of hospitalisation.

## Conclusions

Preliminary clinical trial results suggest a favourable benefit-risk profile for remdesivir compared to placebo, however there is limited safety data available at the current time. The current framework summarises the key anticipated benefits and risks for which further data are needed. Ongoing clinical trial data can be incorporated into the framework when available to provide an updated benefit-risk assessment.

## Data Availability

Data used in this analysis are available from the references supplied.

## Notes

**Conflicts of interest/Competing interests** The Drug Safety Research Unit is an independent charity (No. 327206), which works in association with the University of Portsmouth. It receives unconditional donations from pharmaceutical companies. The companies have no control on the conduct or the publication of the studies conducted by the DSRU. Gilead is providing support for a methodological project led by the DSRU as a part of a large group of pharma companies, unrelated to this product. Miranda Davies, Vicki Osborne, Samantha Lane, Debabrata Roy, Sandeep Dhanda, Alison Evans and Saad Shakir have no conflicts of interest to declare.

### Competing Interest Statement

The Drug Safety Research Unit is an independent charity (No. 327206), which works in association with the University of Portsmouth. It receives unconditional donations from pharmaceutical companies. The companies have no control on the conduct or the publication of the studies conducted by the DSRU. Gilead is providing support for a methodological project led by the DSRU as a part of a large group of pharma companies, unrelated to this product. Miranda Davies, Vicki Osborne, Samantha Lane, Debabrata Roy, Sandeep Dhanda, Alison Evans and Saad Shakir have no conflicts of interest to declare.

### Funding Statement

No funding was received for this project.

